# Neural Basis of Pain Empathy Dysregulations in Mental Disorders – A Pre-registered Neuroimaging Meta-Analysis

**DOI:** 10.1101/2024.02.07.24302440

**Authors:** Jingxian He, Mercy Chepngetich Bore, Heng Jiang, Xianyang Gan, Junjie Wang, Jialin Li, Xiaolei Xu, Lan Wang, Kun Fu, Liyuan Li, Bo Zhou, Keith Kendrick, Benjamin Becker

**Affiliations:** The Center of Psychosomatic Medicine, Sichuan Provincial Center for Mental Health, Sichuan Provincial People’s Hospital, University of Electronic Science and Technology of China, Chengdu, China; School of Life Science and Technology, University of Electronic Science and Technology of China, Chengdu, China; State Key Laboratory of Brain and Cognitive Sciences, The University of Hong Kong, Hong Kong, China; Department of Psychology, The University of Hong Kong, Hong Kong, China; Max Planck School of Cognition, Stephanstrasse 1a, Leipzig, Germany; School of Psychology, Shandong Normal University, Jinan, China

**Keywords:** Empathy, Pain, fMRI, Mental Disorder, Psychiatry

## Abstract

Pain empathy represents a fundamental building block of several social functions, which have been demonstrated to be impaired across various mental disorders by accumulating evidence from case-control functional magnetic resonance imaging (fMRI) studies. However, it remains unclear whether the dysregulations are underpinned by robust neural alterations across mental disorders. This study utilized coordinate-based meta-analyses to quantitatively determine robust markers of altered pain empathy across mental disorders. To support the interpretation of the findings exploratory network-level and behavioral meta-analyses were conducted. The results revealed patients with mental disorders exhibited increased pain empathic reactivity in the left anterior cingulate gyrus, adjacent medial prefrontal cortex, and right middle temporal gyrus, yet decreased activity in the left cerebellum IV/V and left middle occipital gyrus compared to controls. The hyperactive regions showed network-level interactions with the core default mode network (DMN) and were associated with affective and social cognitive domains. The findings suggest that pain-empathic alterations across mental disorders are underpinned by excessive empathic reactivity in brain systems involved in empathic distress and social processes, highlighting a shared therapeutic target to normalize basal social dysfunctions in mental disorders.

## Introduction

Pain empathy, defined as the capacity to understand and share the emotional and sensory aspects of pain felt by another individual (Fitzgibbon et al., 2010), plays a pivotal role in various social functions, including prosocial behavior (to motivate altruistic behavior) (Hartmann et al., 2022), social bonding (foster a sense of trust and emotional connection), and cooperation (Karos et al., 2020; Novembre et al., 2015), as well as adaptive emotion regulation (to improve understanding and expression of emotional experiences) (Fitzgibbon et al., 2010). Moreover, impaired empathy has been associated with a decline in social functioning, which is considered a key feature of psychotic spectrum disorders (Kuis et al., 2021), and across individuals with mental disorders deficits in the cognitive and affective component of empathy may lead to difficulties in interpreting the thoughts and feelings of others (Henry et al., 2008), potentially leading to reduced socialization and increased interpersonal distress (Bailey et al., 2008). Dysfunctions in these domains have been widely observed in common mental disorders (see ref. (Kennedy and Adolphs, 2012)), and these dysfunctions may extend beyond traditional diagnostic categories (see ref. (Cuthbert, 2014)). Emerging evidence from studies involving individuals with mental disorders and high levels of pathology-related traits indicate shared pain empathic dysfunctions (see refs. (Corbera et al., 2014; Li et al., 2019; Meng et al., 2019; Xu et al., 2020; Zhang et al., 2023)), with most studies reporting increased empathic distress in individuals with depression, autism spectrum disorders (ASD), schizophrenia (SCH) and elevated levels of alexithymia (Corbera et al., 2014; Guhn et al., 2020; Horan et al., 2016; Lamm et al., 2016), although results remained inconsistent in ASD (Meng et al., 2019; Zhang et al., 2023). Consequently, as a key capacity supporting social and self-related functions, abnormalities in pain empathy may lead to social deficits as well as increased empathic distress in individuals with mental disorders (Di Girolamo et al., 2022; Thirioux et al., 2019). They may serve as common deficits across several mental disorders and promote functional impairments in daily life.

The neural basis of pain empathy has been extensively examined using functional magnetic resonance imaging (fMRI) in healthy individuals (Fallon et al., 2020; Labek et al., 2023; Lamm et al., 2011; Paradiso et al., 2021). Together with conceptual perspectives, these studies have outlined a core network that underlies pain-empathic processing including the anterior and middle cingulate cortex (ACC, MCC), middle and anterior insula cortex (MIC, AIC) as well as prefrontal and sensorimotor regions (Betti and Aglioti, 2016; Shackman et al., 2011; Timmers et al., 2018; Yao et al., 2016; Zhou et al., 2020b). Within this network, the cingulate and insula primarily extract the affective dimension of pain and interact with the subcortical, motor, and prefrontal regions to initiate adaptive responses (Dixon et al., 2017).

Pain empathy studies in healthy individuals and patient populations used fMRI in combination with different experimental paradigms and stimuli including e.g. visual stimuli presenting noxious stimulation of the limbs as well as painful facial expressions. The stimuli have been extensively characterized in previous studies in terms of the affective experience they induce using rating scales. A direct comparison of stimuli presenting noxious manipulation of limbs and painful facial expressions confirmed that both stimuli induce pain-empathic experiences and arousal (see e.g. (Zhou et al., 2020a)) and engage core brain systems of pain empathy, including the insula and anterior cingulate cortex (Jauniaux et al., 2019). However, stimuli presenting noxious stimulation of the limbs may provide more objective and clear indications of pain experienced by the other person and may lead to a stronger experience of pain empathy and the engagement of inferior frontal brain regions involved in the encoding of sensory information, whereas painful facial expression may require a more subjective appraisal and may lead to a stronger engagement of medial prefrontal and temporal regions involved in social processes (Jauniaux et al., 2019; Zhou et al., 2020a; Zhou et al., 2016). Based on an increasing number of studies reporting pain-empathic deficits in mental disorders - which may encompass both, vicarious and appraisal-related components of empathy (Corbera et al., 2014; Meng et al., 2019; Zhang et al., 2023) - case-control studies combined pain empathy paradigms with fMRI to determine the underlying neurofunctional alterations in depression, SCH, or ASD (Fan et al., 2014; Fujino et al., 2014; Gu et al., 2015; Horan et al., 2016; Vistoli et al., 2017; Xu et al., 2020). However, these studies commonly compared a single group of patients with controls (but see ref. (Xu et al., 2020)) and findings with respect to robust neural alterations remained inconclusive. Several studies reported that the ACC, middle temporal gyrus (MTG), and inferior frontal gyrus (IFG) exhibit stronger pain empathic reactivity in patients (Gu et al., 2015; Hadjikhani et al., 2014; Horan et al., 2016; Ruetgen et al., 2021), while other studies reported no or opposite alterations (Fan et al., 2014; Xu et al., 2020). Moreover, results regarding the AIC - a region crucially involved in pain-empathic experience (Gu et al., 2012; Yao et al., 2016) - remained inconsistent, with several studies reporting intact AIC pain-empathic reactivity in the context of cerebellar alterations in mental disorders (Gu et al., 2015; Hadjikhani et al., 2014; Moriguchi et al., 2007).

The lack of convergent evidence for shared neural alterations may reflect disorder-specific neurofunctional empathy dysregulations or methodological limitations inherent to the conventional case-control neuroimaging approach, including (1) limited sample size, (2) confinement to a single disorder, and (3) variations related to neuroimaging analysis methods (Müller et al., 2018; Specht, 2019; Zhou et al., 2022). Coordinate-based Meta-Analysis (CBMA) can help to overcome these limitations, including small sample sizes or the focus on a single disorder by pooling data from multiple studies, thus enhancing statistical power and generalizability. This method not only consolidates findings across diverse populations and imaging techniques but also identifies consistent brain activation and connectivity patterns, providing robust neural correlates for specific tasks or conditions. CBMA also explores variability between studies, and can thus help to identify inconsistencies and discrepancies (Klugah-Brown et al., 2024; Radua et al., 2012; Radua et al., 2013). Previous meta-analytic studies have successfully employed CBMA to determine common and separable neurofunctional alterations in mental disorders in cognitive, affective, and reward processing domains (e.g., (Bore et al., 2024; Ferraro et al., 2022; Klugah-Brown et al., 2020; McTeague et al., 2017; McTeague et al., 2020)), yet, pain empathic processing alterations across mental disorders have not been systematically explored.

Against this background, the present pre-registered meta-analysis capitalized on the growing number of original case-control fMRI studies on pain empathy and utilized a pre-registered meta-analytic strategy to determine brain regions that exhibit the most robust alterations during pain empathic processing across mental disorders. To further inform and guide the interpretation of the findings we included network-level and behavioral meta-analyses. The network-level analyses were conducted for the regions initially identified in the primary meta-analysis to determine which larger brain networks the identified regions functionally engage with. Behavioral meta-analyses were additionally employed to provide a behavioral characterization of the identified regions in terms of behavioral domains that have most commonly been associated with these regions in the previous literature. Based on the previous literature, we hypothesized shared alterations in core regions of the pain empathic network (insula and ACC) and regions underlying social processing (e.g., regions in the temporal cortex) (Feng et al., 2021; Lamm et al., 2011; Zhou et al., 2020b).

## Methods

### Literature Search

The meta-analysis adhered to the Preferred Reporting Items for Systematic Reviews and Meta-Analyses (PRISMA) guidelines (**Figure 1**) (Page et al., 2021) and was preregistered on the Open Science Framework platform (https://osf.io/axt9k/). Literature search identified original case-control fMRI studies examining neural activation during pain-empathic processing (in response to stimuli showing noxious manipulation to limbs or painful facial expressions, for shared neural and behavioral effects see ref. (Zhou et al., 2020b)) in mental disorders according to the Diagnostic and Statistical Manual of Mental Disorders (Version 5, DSM-5). The search utilized three databases: PubMed, Web of Science, and Scopus. The search period included the period until May 2024.

**Figure 1.**
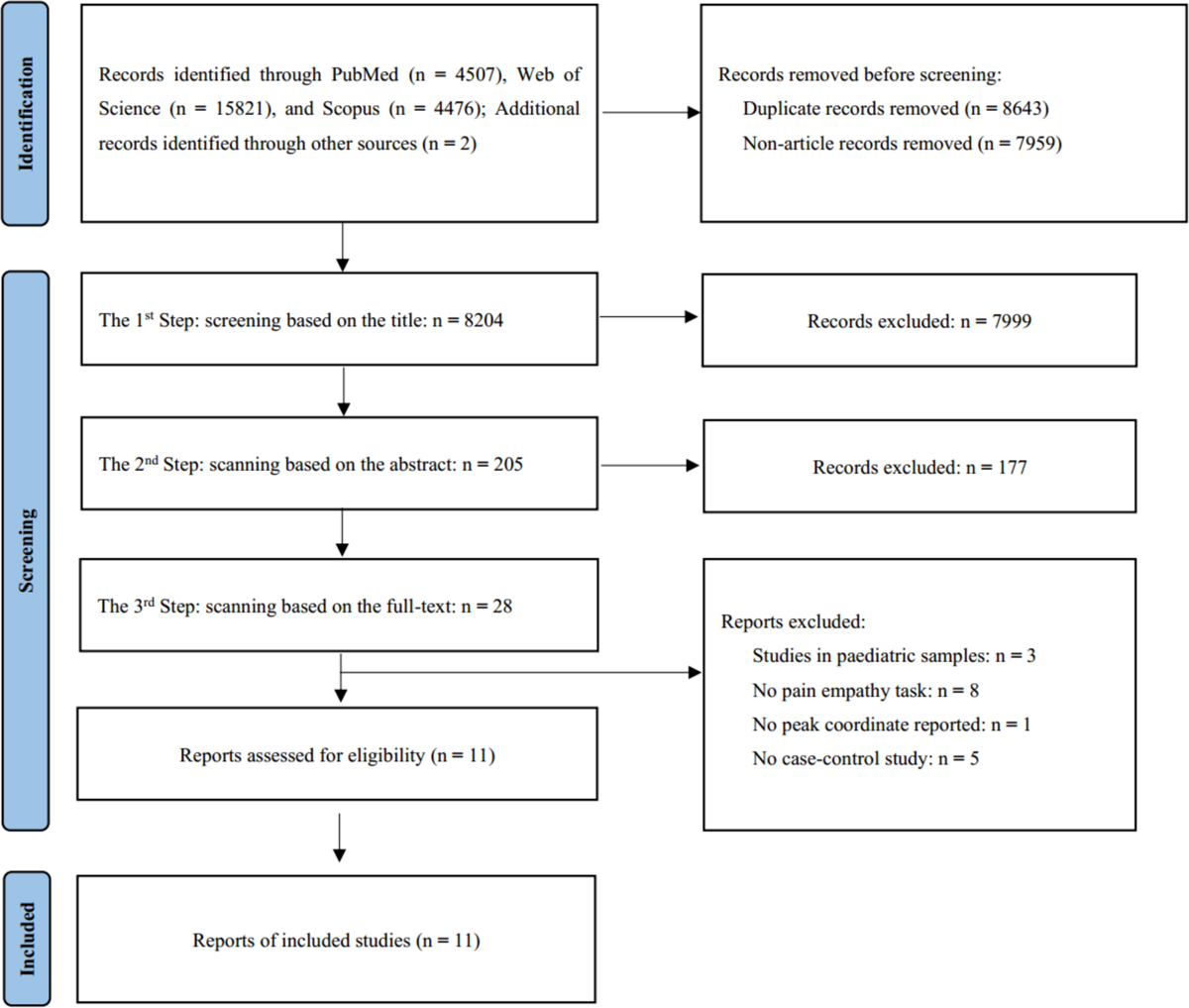
Flowchart of literature search and screening

The search strategies, initially designed for PubMed, combine MeSH (Medical Subject Headings) terms and keywords and were subsequently adapted and utilized with the other two databases (main focus was on pain empathy, functional magnetic resonance imaging, mental disorders; detailed terms, see **Supplement 1**). According to DSM-V criteria, individuals with mental disorders including MDD, SCH, ASD, alexithymia, and CD experience abnormalities in pain empathy causing social deficits as well as empathic distress. Alexithymia is a pathology-relevant personality trait characterized by difficulties in recognizing, expressing, and describing one’s own emotional experiences. Elevated levels of alexithymia represent a common symptom in many psychiatric conditions, including depression and anxiety disorders. In the context of long-standing discussions on the role of alexithymia in pain-empathic alterations, we here included studies in individuals with high levels of alexithymia (see refs. (Brewer et al., 2015; Li et al., 2019)). In addition, we screened published reviews and meta-analyses on related topics to identify further relevant literature.

Two independent reviewers (JX.H. and M.C.B.) assessed inclusion criteria, and conflicts were resolved by a third reviewer (B.B.). Full-context articles were reviewed by two independent reviewers (JX.H. and M.C.B.). Data extraction was performed by two reviewers and conflicts and errors in data extraction were resolved by a third reviewer (B.B.).

### Inclusion and Exclusion Criteria

The initial exclusion of existing articles, including duplicates and non-articles, was first performed using Endnote software (X9 version; https://endnote.com/). We next carefully screened the literature according to standardized criteria and guidelines (**Figure 1**). The following inclusion criteria were applied: (1) studies using a pain empathy task (presenting stimuli depicting painful face expression or body pain to the participants); (2) studies acquiring and analyzing whole brain data acquisition (excluding regions of interest [ROI] results); (3) original studies utilizing a case-control design comparing brain activity in patients and healthy individuals; (4) activation contrasts generated by the subtractive approach reflecting differences between patients and controls during the processing of pain empathy; (5) differences in terms of peak brain activation coordinates reported; (6) results reported in standard stereotactic coordinates (either Talairach or Montreal Neurological Institute [MNI] space), and (7) article written in English.

Additional exclusion criteria were as follows: (1) studies in pediatric samples; (2) studies reporting results without exact peak coordinates; (3) studies that focus on other empathy domains such as affective or cognitive empathy; (4) studies reporting results in patients or healthy subjects only; (5) studies reporting findings from datasets from previous (already included) studies.

### Voxel-Wise Meta-Analysis

The meta-analysis was performed using Seed-based d Mapping Permutation of Subject Images (SDM-PSI) (version 6.22, https://www.sdmproject.com). SDM combines recent advances and features of previous methods, such as activation likelihood estimate (ALE) and multilevel kernel density analysis (MKDA), and permits more detailed and accurate meta-analyses (Radua and Mataix-Cols, 2018; Radua et al., 2012). SDM-PSI is based on the MetaNSUE algorithm using unbiased maximum likelihood estimation (MLE) to build whole-brain effect size and variance maps, while taking into account additions and subtractions of measurements (e.g., activation and de-activation) that allow conflicting results to cancel each other out. The procedure moreover takes into account irregular local spatial covariances of different brain tissues. Meanwhile, SDM-PSI uses a leave-one-out jackknife procedure, weighting the calculation of within-study variance (i.e., studies with larger sample sizes and/or lower errors contribute more) and between-study heterogeneity, to prevent a single study or few studies from driving the results. Although image-based meta-analysis can offer more precise results, the scarcity of original studies and authors providing original t-maps limits its practical application (Liu et al., 2022; Salimi-Khorshidi et al., 2009). For pooling of data with SDM-PSI, the coordinates in standard spaces and their respective *t*-values are collected. An anisotropic Gaussian kernel with a full-width at half maximum (FWHM) of 20*mm* and a voxel size of 2*mm* was used (Albajes-Eizagirre et al., 2019). To balance type I and type II errors when determining neurofunctional brain alterations in mental disorders results were thresholded at *p* < 0.0025, uncorrected; *k* > 10 (in line with (Bore et al., 2023; Chavanne and Robinson, 2021)) (Radua et al., 2012). Given that mental disorders have different onset and peak ages (Solmi et al., 2021) and neurodevelopmental changes have been reported in the domain of pain empathy (Decety and Michalska, 2010), age was included as a covariate (in line with (Gong et al., 2020)), to further account for a potential impact of the variations in age between the samples in the included studies we employed an independent sample t-test by SPSS to the standard deviations for age reported in the original studies. Results did not confirm significant differences in the age variations between patients and controls (*df* = 20, *t-value* = 0.13, *p* = 0.686). Visualization of brain activation was performed using MRIcroGL (version 1.2.20, https://www.nitrc.org/projects/mricrogl).

### Heterogeneity and Bias Testing

Heterogeneity in the present study was assessed using the *I*^2^ index, with *I*^2^ values of 25%, 50%, and 75% representing low, moderate, and high heterogeneity respectively (Higgins, 2002). Tests were carried out to check for publication bias. Egger’s test was used to assess funnel plot asymmetry, with a *p*-value less than 0.05 indicating significant publication bias (Egger et al., 1997).

### Functional Characterization of the Identified Regions at the Network and Behavioral Level

To investigate the brain networks associated with the identified regions, peak coordinates of the identified regions were entered into the Neurosynth database (https://www.neurosynth.org) to obtain non-thresholded resting-state functional connectivity maps (rs-FC maps) and meta-analytic co-activation maps of the relevant brain regions. Subsequently, we employed SPM12 (Welcome Department of Imaging Neuroscience, London, UK) to apply a threshold of *r* > 0.2 to the rs-FC maps and convert the thresholded maps into *Z*-scores. Finally, we merged the rs-FC thresholded maps with the meta-analytic co-activation maps to generate conjunction maps of the relevant regions of interest. Next, we employed the Anatomy toolbox to extract the labels of the conjunction maps (Eickhoff et al., 2005).

For further behavioral characterization of the identified regions, we implemented meta-analytic behavioral profiling using the Brain Annotation Toolbox (BAT) in combination with AAL2 (Automated Anatomical Labeling, atlas). To this end, the peak coordinates of the identified regions were examined and the top 15 behavioral items and *p*-values were obtained (in descending order of *p*-values) (Liu et al., 2019).

### Meta-Regression Analyses

Meta-regression analysis was employed to assess the effect of subject characteristics on the obtained results, including gender, age, and treatment. Significant findings suggest that certain variables are related to the observed heterogeneity, indicating the potential influence of unaccounted factors or confounding variables.

## Results

### Characteristics of the Included Studies

Based on the standardized criteria and guidelines, we finally included 11 studies from 525 individuals (Decety et al., 2009; Fan et al., 2014; Fujino et al., 2014; Gu et al., 2015; Hadjikhani et al., 2014; Horan et al., 2016; Lockwood et al., 2013; Moriguchi et al., 2007; Ruetgen et al., 2021; Vistoli et al., 2017; Xu et al., 2020), 296 patients and 229 controls, mean age 28.2 ± 9.1 years, and one on conduct disorder (CD), two on alexithymia, two on SCH, three on ASD, and three on major depressive disorder (MDD) (details in **Table 1**). We exported 143 peak coordinates on pain empathy differences between the comparisons of patient and control group, including 8 physical pain empathy paradigms, and 3 affective pain empathy (face expression) paradigms (details in **Table 2**).

**Table 1.** Demographic characteristics of the 11 fMRI studies included in the meta-analysis.

| Study ID | Disease type | Num. | Female (%) | Patient |  |  | Control |  |
| --- | --- | --- | --- | --- | --- | --- | --- | --- |
|  |  |  |  | n<sub>1</sub> | Mean age (SD) | Treatment (Y/N) | n<sub>2</sub> | Mean age (SD) |
| Bird et al, 2010 | Alexithymia | 36 | 0% | 18 | 34.6(13.3) | N | 18 | 35(12.8) |
| Decety et al, 2009 | Conduct Disorder (CD) | 38 | 0% | 30 | 17(NA) | N | 8 | 17(NA) |
| Fan et al, 2013 | Autism Spectrum Disorder (ASD) | 45 | 0% | 24 | 18.4(2.8) | N | 21 | 19.3(3.4) |
| Fujino et al, 2014 | Major Depressive Disorder (MDD) | 22 | 77% | 11 | 37.2(9.5) | Y | 11 | 32.3(10.5) |
| Gu et al, 2015 | Autism Spectrum Disorder (ASD) | 29 | 0% | 15 | 26.2(6.4) | N | 14 | 26.8(7.8) |
| Hadjikhani et al, 2014 | Autism Spectrum Disorder (ASD) | 67 | 8% | 36 | 23.5(8.7) | N | 31 | 22.5(7.5) |
| Horan et al, 2016 | Schizophrenia | 42 | 31% | 21 | 48.2(10.4) | Y | 21 | 46.5(7.1) |
| Moriguchi et al, 2007 | Alexithymia | 30 | 83% | 16 | 20.2(1) | N | 14 | 20.4(0.94) |
| Ruetgen et al, 2021 | Major Depressive Disorder (MDD) | 96 | 70% | 61 | 27.34(1.35) | Y | 35 | 27.41(1.3) |
| Vistoli et al, 2017 | Schizophrenia | 48 | 17% | 27 | 29.7(8.6) | N | 21 | 29.2(7.9) |
| Xu et al, 2020 | Major Depressive Disorder (MDD) | 72 | 0% | 37 | NA | N | 35 | NA |
| Abbreviation: Num, total number of subjects; n<sub>1</sub>, number of patients; n<sub>2</sub>, number of controls; Treatment, the patient whether received clinical treatment or not; Y, received treatment; N, not received treatment; NA, not available. |  |  |  |  |  |  |  |  |

**Table 2.** Data characteristics of the 11 fMRI studies included in the meta-analysis.

| Study ID | Correction | Space | Coordinates (Pain > No Pain)<sup>a</sup> | Paradigm<sup>b</sup> |
| --- | --- | --- | --- | --- |
| Bird et al, 2010 | p<0.001 uncorrected | MNI | HC > Alexithymia | Physical Pain |
| Decety et al, 2009 | p<0.005 uncorrected | MNI | CD > HC | Physical Pain |
| Fan et al, 2013 | p<0.05 corrected | MNI | HC > ASD | Physical Pain |
| Fujino et al, 2014 | p<0.001 uncorrected | Talairach | MDD > HC & HC > MDD | Physical Pain |
| Gu et al, 2015 | p<0.05 uncorrected | MNI | ASD > HC & HC > ASD | Physical Pain |
| Hadjikhani et al, 2014 | p<0.01 uncorrected | MNI | ASD > HC & HC > ASD | Expression Pain |
| Horan et al, 2016 | p<0.05 corrected | MNI | SCH > HC & HC > SCH | Expression Pain |
| Moriguchi et al, 2007 | p<0.005 uncorrected | MNI | Alexithymia > HC & HC > Alexithymia | Physical Pain |
| Ruetgen et al, 2021 | p<0.05 corrected | MNI | MDD > HC & HC > MDD | Expression Pain |
| Vistoli et al, 2017 | p < 0.001 uncorrected | MNI | HC > SCH | Physical Pain |
| Xu et al, 2020 | p<0.05 corrected | MNI | HC > MDD | Physical Pain |
| Abbreviation: HC, Healthy Control; CD, Conduct Disorder; ASD, Autism Spectrum Disorder; SCH, Schizophrenia; MDD, Major Depressive Disorder; MNI, Montreal Neurological Institute space.<br><sup>a</sup> Pain Condition: participants view images of individuals undergoing pain or injury; No Pain Condition: participants are shown neutral images devoid of pain or harm, functioning as a control; The greater than symbol (>) signifies that peak coordinate activation is higher in one group compared to another.<br><sup>b</sup> Physical Pain: stimuli involve actual bodily injury or situations that suggest physical pain, prompting participants to perceive or envision the sensory experience of pain. Expression Pain: emphasizes stimuli displaying facial expressions of pain, with a focus on the emotional reaction to observed distress in others. |  |  |  |  |

### Shared Pain Empathy Alterations across Mental Disorders

The primary meta-analysis revealed that patients with mental disorders exhibited increased pain empathic activation in a cluster located in the left ACC (x/y/z = 0/48/0, *p* < 0.0025) and spreading into the medial prefrontal cortex (mPFC) and a second cluster located in the right MTG (x/y/z = 60/-14/-12, *p* < 0.0025) in comparison with controls (**Table 3**, **Figure 2A**). Reduced activation in the patients relative to controls was observed in the left cerebellum Ⅳ/Ⅴ (CE_4/5, x/y/z = −22/-48/-28, *p* < 0.0025) and the left middle occipital gyrus (MOG, x/y/z = - 52/-70/2, *p* < 0.0025) (**Table 3**, **Figure 2B**). Heterogeneity test results were all below 25%, indicating low heterogeneity across studies. Additionally, the results of Egger’s test revealed *p*-values greater than 0.05, suggesting that the included studies did not demonstrate evidence of a publication bias (**Table 3**).

**Figure 2.**
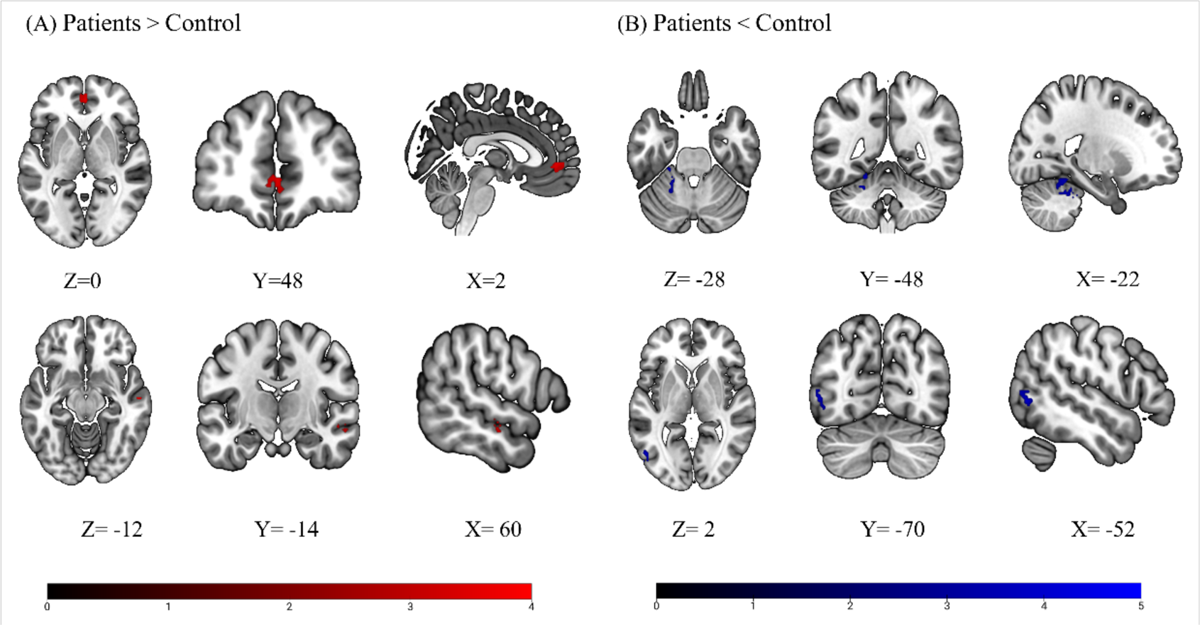
Meta-analytically determined neurofunctional alterations during pain empathy in patients with mental disorders (A) Compared to controls, patients with mental disorders exhibited increased activation in the left anterior cingulate gyrus (activation spreading to the medial prefrontal cortex) and the right middle temporal gyrus. (B) Patients with mental disorders showed less activation in the left cerebellar area Ⅳ/Ⅴ and the left middle occipital gyrus compared to controls. Results displayed at *p* < 0.0025, uncorrected; *k* > 10, coordinates reported in Montreal Neurological Institute (MNI) space.

**Table 3.** Detailed meta-analytic results, heterogeneity tests, and publication bias.

| Comparison | Cluster | Voxel | MNI coordinates(x,y,z)<sup>a</sup> | SDM-Z | Voxel $P^b$ | Hedges'g | $I^2$ | Egger test $P^c$ |
| --- | --- | --- | --- | --- | --- | --- | --- | --- |
| Patient > Control | Anterior Cingulate, Left (ACC.L) | 69 | 0,48,0 | 3.510 | 0.000224233 | 0.392553 | 0.355083 | 0.931 |
|  | Middle Temporal Gyrus, Right (MTG.R) | 17 | 60, -14, -12 | 3.699 | 0.000108063 | 0.414338 | 0.276461 | 0.996 |
| Patient < Control | Cerebellum IV/V, Left (CE_4_5) | 108 | -22, -48, -28 | -3.797 | 0.000073254 | -0.466374 | 2.347702 | 0.960 |
|  | Middle Occipital Gyrus, Left (MOG.L) | 52 | -52, -70,2 | -4.246 | 0.000010848 | -0.490509 | 0 | 0.920 |
| <sup>a</sup> MNI = Montreal Neurological Institute space<br><sup>b</sup> Voxel $p < 0.0025$ (uncorrected), significant<br><sup>c</sup> Egger test $p < 0.05$, significant. | | | | | | | | |

### Network-Level Functional Characterization via Neurosynth

The identified ACC region showed strong and overlapping co-activation and connectivity patterns with cortical midline regions including ACC, medial and orbitofrontal regions as well as the posterior cingulate (PCC) and precuneus (**Figure 3A**). The identified MTG region exhibited overlapping co-activation and connectivity patterns with bilateral middle temporal and (inferior) parietal regions as well as with cortical midline regions, i.e., medial prefrontal/cingulate regions and PCC and precuneus (**Figure 3B**). Examining the overlap between both networks revealed that both regions exhibited functional interactions with the cortical midline regions, including the orbitofrontal and medial PFC as well as posterior parietal regions (angular gyrus, PCC, precuneus, **Figure 3C**).

**Figure 3.**
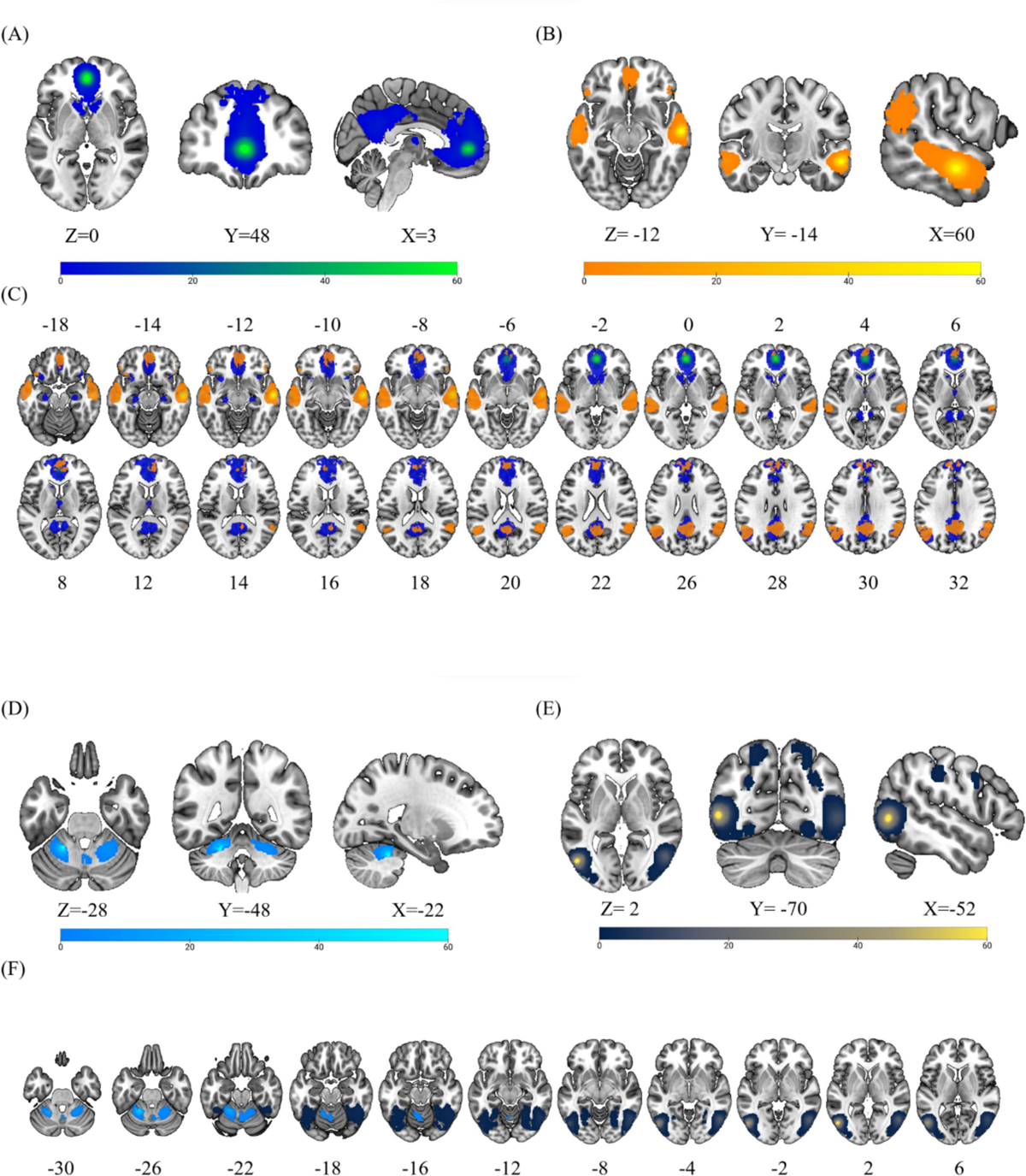
Conjunction maps between the meta-analytic resting state (intrinsic) connectivity maps and the meta-analytic co-activation (task state) maps of regions that exhibited increased and decreased pain empathic reactivity in patients with mental disorders. (A) Conjunction maps for the identified anterior cingulate region (with peak coordinates at MNI 0, 48, 0). (B) Conjunction maps for the identified middle temporal gyrus region (with peak coordinates at MNI 60, −14, −12). (C) Overlap of the two large-scale networks identified in A and B (conjunction maps). (D) Conjunction maps for the identified cerebellum region (with peak coordinates at MNI −22, −48, −28). (E) Conjunction maps for the identified middle occipital gyrus region (with peak coordinates at MNI −52, −70, 2). (F) Overlap of the two large-scale networks identified in D and E (conjunction maps). Coordinates are in the MNI space.

Examination of the connectivity and co-activation maps of the regions exhibiting decreased pain empathic reactivity in mental disorders (cerebellum, MOG) revealed separable networks. The cerebellum region exhibited regional-restricted bilateral connectivity with other (anterior) cerebellar regions (**Figure 3D**), while the MOG exhibited interactions with a bilateral network encompassing lateral occipital regions, and precentral and postcentral regions (**Figure 3E**).

### Behavioral Characterization of the Identified Regions

Meta-analytic behavioral characterization indicated that the left ACC and right MTG are associated with negative affective (stress, depression, fear) and evaluative (self-referential, reward) processes or social cognitive (theory of mind, social) processes, respectively (**Figure 4A, B**). In contrast, the left CE 4/5 and left MOG were found to be associated with motor and visuospatial processes (**Figure 4C, D**). Please note that smaller *p*-values indicate a higher degree of correlation with the relevant brain regions.

**Figure 4.**
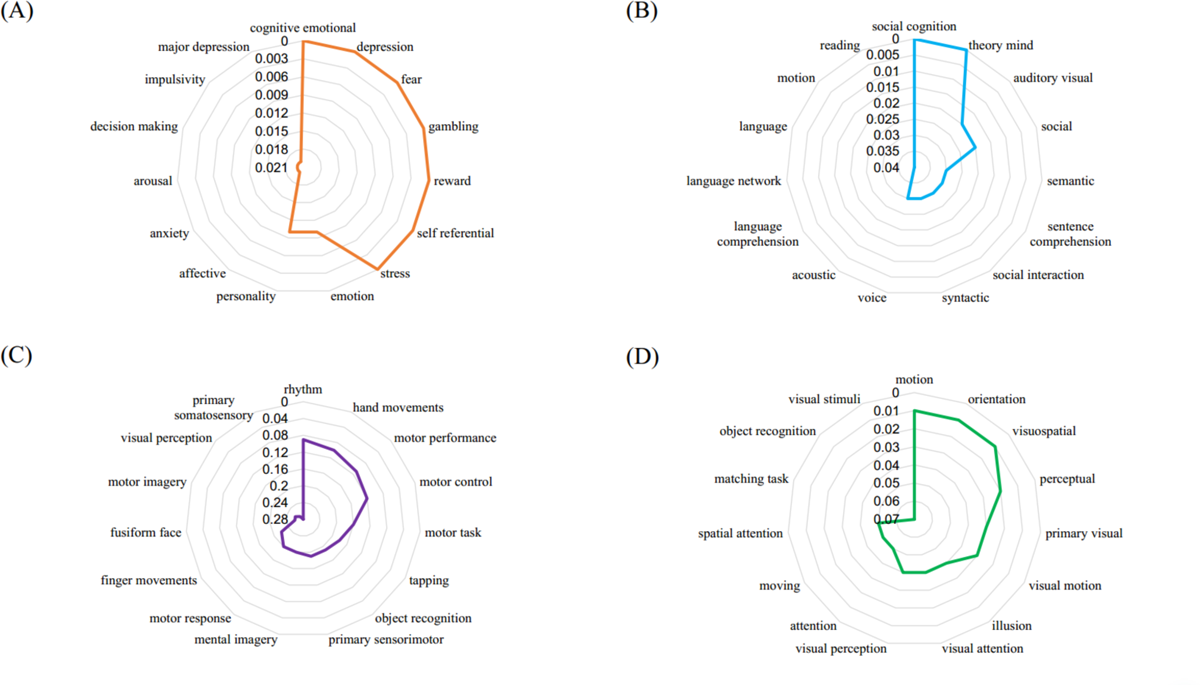
Radar chart of the top 15 behavioral terms associated with the four peak coordinates. (A) Left anterior cingulate gyrus and relevant behavioral terms. (B) Right middle temporal gyrus and relevant behavioral terms. (C) Left cerebellar Ⅳ/Ⅴ and relevant behavioral terms. (D) Left middle occipital gyrus and relevant behavioral terms. Please note that a smaller *p*-value indicates a higher degree of correlation with the relevant brain regions.

### Meta-Regression Analyses: Gender, Age, and Treatment

Meta-regression analysis in SDM revealed no significant effects of gender, age, or medication on altered activity in the four meta-analytically identified brain regions.

## Discussion

We capitalized on pre-registered neuroimaging meta-analyses to determine whether pain-empathic dysregulations share a common neurobiological substrate across mental disorders. To guide interpretation, the primary analyses were flanked by meta-analytic network-level and behavioral profiling of the identified regions. Quantitative analysis of eleven case-control fMRI studies with data from 296 patients and 229 controls revealed that individuals with mental disorders exhibited increased pain empathic reactivity in the left ventral ACC and adjacent mPFC, as well as in the contralateral MTG. Conversely, patients with mental disorders showed hyporeactivity in the left anterior cerebellum and MOG. Network-level decoding analyses further indicated that the hyperactive regions show a strong and convergent network-level interaction with cortical midline regions that together constitute the core default mode network (DMN) and were behaviorally characterized by stress, negative affect, and social cognition-related terms. In contrast, the hypoactive regions exhibited distinct and regional-specific connectivity profiles with the anterior cerebellum or occipital and sensory-motor regions, respectively. With reference to our hypotheses, these findings underscore that shared neural dysregulations in the ACC and MTG may underlie pain-empathic alterations across mental disorders. In contrast to our hypothesis, no alterations in the insular cortex were observed, while the anterior cerebellum and MOG showed convergent hyporeactivity.

### Shared Pain Empathic Neural Reactivity across Mental Disorders

Animal models suggest that the ACC plays a critical role in both, the experience and regulation of first-hand experience of pain and negative affective states as well as social transmission of pain and distress (see refs. (Carrillo et al., 2019; Li et al., 2021; Smith et al., 2021)). It represents the core module for the affective-motivational component of the “pain matrix” activated by various painful stimuli (Duerden and Albanese, 2013) and plays an important role in the transition from acute to chronic pain (Bliss et al., 2016; Kuner and Kuner, 2021; Li et al., 2023). In human imaging studies, the ACC and insula have been consistently determined as core regions of the pain empathic network, yet most meta-analyses identified a more dorsal part of the ACC (see ref. (Lamm et al., 2011)). In contrast to the dorsal part of the ACC, the ventral ACC (vACC) has been strongly involved in evaluating the emotional salience of a situation and to adaptively regulate the emotional experience (see refs. (Etkin et al., 2011a; Xiang et al., 2018)). Hyperactivity in this region may thus reflect a stronger distress-related pain-empathic reactivity or a stronger need to regulate the negative affective experience (Betti and Aglioti, 2016; Nabulsi et al., 2022). An increasing number of reports indicated that individuals with mental disorders experience greater levels of empathic distress (Allman et al., 2001; Apps et al., 2016) and the role of this region in the experience of pain-related emotional distress may reflect higher pain empathic reactivity in individuals with mental diorders. The vACC subregion of the ACC has moreover been found to play an important role in other related functions such as social cognitive decision-making, behavior encoding, and outcome prediction (Brown and Morey, 2012; Lockwood and Wittmann, 2018) and can mediate the saliency of the external stimuli thus triggering pain empathic experiences (Lamm et al., 2016).

Further mapping the location of the significant peak coordinates with the Juelich Brain atlas revealed that the ACC cluster was located specifically in the pregenual subdivision (pgACC). The pgACC has been associated with several functions that support pain empathic processing, including pain empathic reactivity and habituation (Preis et al., 2015), compassion (Klimecki et al., 2014) and implicit emotion regulation (Etkin et al., 2010). The cluster extended into the mPFC, an integral component of the DMN, a network critically engaged in several functions supporting pain empathy, i.e., socio-affective processes such as self-reference, social cognition, and contextual integration (see refs. (Menon, 2023; Yeshurun et al., 2021)).

The ACC cluster also encompassed regions of the mPFC, a region with strong anatomical and functional connections with a wide range of subcortical brain regions (Ong et al., 2019). Both animal and human studies have underscored the significance of the ACC and the mPFC in emotional cognitive processing, social-emotional cognition (e.g., attachment, empathy, pro-social behavior), emotion regulation, decision-making, and pain (Etkin et al., 2011b; Jahn et al., 2016; Klein-Flügge et al., 2022; Malezieux et al., 2023; Murray and Fellows, 2021; Zhou et al., 2019; Zhuang et al., 2021). The convergent dysfunction affecting both of these systems aligns with the additional functional requirements of pain empathy, which, compared to the experience of direct pain require complex social-emotional computations to determine the experience of the other person and the integration of previous knowledge and contextual information (Betti and Aglioti, 2016; Danziger et al., 2009). In line with this suggestion, previous studies reported that structural damage or aberrant functional changes in the ACC or mPFC can severely impair social behavior as well as adaptive emotion regulation in animals (Bissière et al., 2008; Bliss-Moreau et al., 2021; Chudasama et al., 2013; Kaefer et al., 2020; Ohta et al., 2020).

In addition to the ACC/mPFC, the MTG exhibited consistently elevated pain empathic reactivity across mental disorders. The MTG has been convergently involved in social-cognitive functions that may support the pain-empathic process, including inferring and understanding the psychological states of others (Theory of Mind, ToM, (Preckel et al., 2018; Schurz et al., 2017; Vucurovic et al., 2023)), contextual and sensory information integration (Xu et al., 2019), as well as pain perception and pain empathy (Kucyi and Davis, 2015; Maliske et al., 2023). In this context, a stronger MTG recruitment may reflect an impaired ability to distinguish own and others’ distressing states (Hoffmann et al., 2016; Preckel et al., 2018), leading to an impaired ability to detach from the pain-empathic situation and in turn compromised social cognition (Sprooten et al., 2017; Zhang et al., 2016).

Although the exact functional interaction between the MTG and the mPFC during pain empathic processing remains to be determined both regions form integral core components of the DMN which supports large scale functional interaction between these regions during pain empathy processes. The MTG modulates attentional resources to pain cues (Ballotta et al., 2018; Lee et al., 2021; Marek and Dosenbach, 2018) and can contribute to the integration and regulation of pain sensation and pain emotion together with the ACC in pain processing (Ou et al., 2024).”

Abnormal pain empathy has been associated to further impairments in mental disorders. Dysregulations in pain empathy may lead to hyperlinking of painful stimuli to related experiences in individuals with mental disorders (Fitzgibbon et al., 2010), difficulty in shifting attention (Li et al., 2020), inability to correctly differentiate between feelings of self and others (Preckel et al., 2018), and can increase the experience of distress while reducing the inhibition of negative emotions (Rütgen et al., 2015; Thirioux et al., 2019).

Together, the pattern of hyperactivation of the ACC, mPFC, and MTG in patients with mental disorders and a convergent coupling of these regions with the DMN may reflect that excessive pain empathic distress responses and a failure to regulate these in the context of social processing dysfunctions may constitute a shared dysregulation mediating pain empathic deficits.

### Abnormal Sensorimotor Functioning in Pain Empathy in Individuals with Mental Disorders

The cerebellum and MOG displayed decreased activity to pain empathic engagement across disorders. In line with our meta-analytic behavioral decoding, numerous previous studies have involved these regions in sensory and motor processes. The cerebellum is anatomically connected to the somatosensory cortex and has a similar topographic map of the body, which is considered to play a crucial role in somatosensory function and motor control (Opie et al., 2022; Wiestler et al., 2011). While we did not hypothesize alterations in these regions, some previous studies reported neurofunctional pain empathic alterations in these regions in autism spectrum disorders and attention deficit hyperactivity disorder (ADHD) indicating abnormalities in cerebellar organization and impaired motor executive control (Betti and Aglioti, 2016; Clark et al., 2021). The MOG is considered an important brain area for visual stimulus feature extraction and recognition, as well as body part perception (Thiebaut de Schotten et al., 2014), and reduced activation of the MOG in patients with MDD and ASD has previously been interpreted to reflect impaired visual information processing abilities (Matsuoka et al., 2020; Teng et al., 2018). Together with an early study suggesting that pain empathy-related cerebellar activation may be related to sensory-motor representation and contribute to cognitive understanding of promoting pain empathy (Strick et al., 2009), these results may reflect early sensory-motor processing dysfunction in the patients. These patients may have abnormal perception and processing of pain empathic stimuli which may lead to a failure to accurately perceive and react to social information.

The present meta-analytic study focuses on abnormal activation patterns shared by pain empathy induced by the included paradigms in mental disorders, and included paradigms with spatial similarities in brain activation in other neuroscience studies, such as activation of the MTG, mPFC, ACC, and other relevant brain regions in Zhou et al.’s study on alternative pain (Zhou et al., 2020b); Jauniaux et al.’s meta-analytic study on pain empathy yielded similar results for ACC activation (Jauniaux et al., 2019). Differences in pain empathy, such as empathic pain responses to different pain empathy stimuli and their specific brain activation patterns, could be further explored in the future.

### Limitations

The study has limitations that should be considered. Firstly, the number of studies on pain empathic processing using fMRI in patients with mental disorders in pain empathy was moderate, meta-analyses depend on the selection and quality of the original studies. Several of the studies included in the present meta-analyses incorporated comparably small sample sizes which may in turn limit the statistical testing power of the meta-analyses (Cremers et al., 2017). While the approach to assign weights based on single studies with the sample size towards each study in SDM may – to a certain extent - account for a bias towards findings based on small sample sizes the results of the present meta-analyses need to be interpreted cautiously and the robustness of the findings remain to be re-evaluated with the accumulation of a larger number of original studies. Secondly, the number of studies did not allow to employ sub-group analyses, e.g., with respect to larger symptomatic groups such as internalizing or externalizing disorders which have shown differential neural signatures (see ref. (Yu et al., 2023)).

## Conclusion

The present study suggests that pain-empathic alterations across disorders are neurally underpinned by increased vACC/mPFC and MTG reactivity and decreased cerebellum and MOG reactivity in patients with mental disorders. This pattern may reflect dysregulated affective responses and social deficits that are further reflected in strong network-level interactions of these regions with the DMN. Targeting pain-empathic dysfunctions in mental disorders with tailored behavioral interventions or via brain modulation techniques that can enhance pain empathy (Geng et al., 2018; Yao et al., 2016) may lead to beneficial effects on social cognitive deficits in mental disorders.

## Data Availability

Data available: Yes
How to access data: The data will be uploaded as an Excel file to the Open Science Framework (https://osf.io/axt9k/)
When available: With publication
Who can access: Anyone

## Author contributions

*Concept and design:* Jingxian He, Benjamin Becker.

*Acquisition, analysis, or interpretation of data:* Jingxian He, Mercy Chepngetich Bore, Heng Jiang, Xianyang Gan, Junjie Wang; Jialin Li, Xiaolei Xu, Lan Wang, Kun Fu, Benjamin Becker.

*Drafting of the manuscript:* Jingxian He, Benjamin Becker.

*Critical review of the manuscript for important intellectual content:* Mercy Chepngetich Bore, Heng Jiang, Xianyang Gan, Keith Maurice Kendrick, Benjamin Becker.

*Statistical analysis:* Jingxian He, Mercy Chepngetich Bore.

*Administrative, technical, or material support:* Mercy Chepngetich Bore, Xianyang Gan, Jialin Li, Xiaolei Xu, Liyuan Li, Bo Zhou, Keith Maurice Kendrick.

*Supervision:* Benjamin Becker.

We would also like to thank Dr. Zhenjiang Li from the University of Chinese Academy of Sciences, for his assistance and insights into the manuscript.

## Role of the Funder/Sponsor

The funders had no role in the design and conduct of the study; collection, management, analysis, and interpretation of the data; preparation, review, or approval of the manuscript; and decision to submit the manuscript for publication. Any opinions, findings, conclusions, or recommendations expressed in this publication do not reflect the views of the Government of the Hong Kong Special Administrative Region or the Innovation and Technology Commission.

**Data Sharing Statement: See Supplement 2**

