## Supplement 1. The specific search terms for "Neural Basis of Pain Empathy Dysregulations in Mental Disorders – A Pre-registered Neuroimaging Meta-Analysis" (from another version of this preprint: version 2, 2024-05-14)

(“pain” OR “vicarious” OR “empathy” OR “empathic”) AND (“fMRI” OR “MRI” OR “magnetic resonance imaging” OR “imaging”) AND (“Attention Deficit Hyperactivity Disorder” OR “ADHD” OR “Autism Spectrum Disorder” OR “Autism” OR “Conduct Disorder” OR “Disruptive Mood Dysregulation Disorder” OR “Eating Disorders” OR “Gender Dysphoria” OR “Intellectual Disability” OR “Internet Gaming Disorder” OR “Major Depressive Disorder” OR “Depression” OR “the Bereavement Exclusion” OR “Mild Neurocognitive Disorder” OR “Obsessive-Compulsive and Related Disorders” OR “Obsessive” OR “Paraphilic Disorder” OR “Personality Disorder” OR “Posttraumatic Stress Disorder” OR “PTSD” OR “Schizophrenia” OR “Sleep-Wake Disorders” OR “Specific Learning Disorder” OR “Social Communication Disorder” OR “Somatic Symptom Disorder” OR “Substance-Related and Addictive Disorders” OR “substance use” OR “Addiction” OR “Anxiety” OR “Psychotic” OR “panic disorder” OR “mood disorder” OR “bipolar disorder” OR “alexithymia”).
